# Effect of a low-intensity digital intervention on recovery from depression among older adults in Brazil: a randomised clinical trial

**DOI:** 10.1101/2023.08.08.23293831

**Authors:** Marcia Scazufca, Carina Akemi Nakamura, Nadine Seward, Thiago Vinicius Nadaleto Didone, Felipe Azevedo Moretti, Luara Aragoni Pereira, Marcelo Oliveira da Costa, Caio Hudson Queiroz de Souza, Gabriel Macias de Oliveira, Mariana Mendes de Sá Martins, Monica Souza dos Santos, Pepijn van de Ven, William Hollingworth, Tim J. Peters, Ricardo Araya

**Affiliations:** Instituto de Psiquiatria, Hospital das Clinicas HCFMUSP, Faculdade de Medicina, Universidade de Sao Paulo, Sao Paulo, Brazil; Departamento de Psiquiatria, Faculdade de Medicina FMUSP, Universidade de Sao Paulo, Sao Paulo, Brazil; Health Service and Population Research, Institute of Psychiatry, Psychology and Neuroscience, King’s College London, London, UK; Health Research Institute, University of Limerick, Limerick, Ireland; Health Economics Bristol, Population Health Sciences, Bristol Medical School, University of Bristol, Bristol, UK; Population Health Sciences, Bristol Medical School, and Bristol Dental School, University of Bristol, Bristol, UK

## Abstract

Importance: There is an urgent need to provide scalable solutions to treat depression amongst older adults in poorly resourced settings.

Objective: To assess the effectiveness of the Viva Vida digital psychosocial intervention for the treatment of depression among older adults.

Design: Pragmatic, two-arm, individually randomised controlled trial with a 1:1 allocation ratio.

Setting: Twenty-four primary care clinics in Guarulhos, Brazil.

Participants: Older adults (60+ years) registered with these clinics were contacted by phone for a screening assessment provided that an active mobile number was available from their primary care records. A two-stage screening for depressive symptomatology with the Patient Health Questionnaire (PHQ) two-items followed by PHQ 9-items was applied. Those who scored ≥ 10 on the PHQ-9 were assessed at baseline and invited to participate. A total of 603 participants were recruited between September 2021 and April 2022, and followed-up to September 2022.

Interventions: The Viva Vida psychosocial programme was offered to the intervention arm (n=298). Over six weeks, participants received 48 audio and visual messages based on psychoeducation and behavioural activation sent by WhatsApp. Health professionals were not involved. The control arm received a single message with information about depression (n=305). Both groups received routine primary care.

Main outcomes: The primary outcome was recovery from depression (PHQ-9 score<10) at the three-month follow-up. Depression at five months was a secondary outcome.

Results: Of 603 participants (mean age, 65.1 years; 451 (74.8%) women), 527 (87.4%) completed the three-month follow-up assessment. At this follow-up, 109 of 257 (42.4%) participants in the intervention arm had recovered from depression, compared with 87 of 270 (32.2%) participants in the control arm (adjusted odds ratio: 1.56; 95% CI: 1.07, 2.27; *P*=.021). This benefit was not maintained at the five-month follow-up (adjusted odds ratio: 1.02; 95% CI: 0.71, 1.47; *P*=.892).

Conclusions and relevance: These results demonstrate the usefulness in reducing depressive symptoms using a self-help intervention that can be readily integrated into primary care programmes for treating older adults with depression. More research is needed to understand how the intervention can be optimised to maintain benefits in the longer term.

Trial Registration: ReBEC registration: RBR-4c94dtn (https://ensaiosclinicos.gov.br/rg/RBR-4c94dtn).

**Key points:** Question: Can a low-intensity digital (self-help) psychosocial intervention delivered by automated WhatsApp audio and visual messages over six weeks improve depression recovery among older adults in Brazil?

Findings: In this randomised controlled trial involving 603 older adults with depressive symptomatology, the Viva Vida digital intervention substantially improved recovery from depression compared with enhanced usual care at three months (42.4% versus 32.2%; odds ratio: 1.56). However, this difference was not maintained at five months.

Meaning: The Viva Vida programme is a simple and easily scalable strategy, that can be integrated as a first step in the treatment of older adults with depression in primary care.

## Introduction

Reducing the burden of depression in older adults is a global health priority and is key to ensuring healthy ageing and promoting wellbeing, especially in low and middle-income countries (LMICs) where 69% of the world population aged 65+ years live.^1^ In the last decade, Brazil’s number of older adults (60+ years) increased by 40%, corresponding to 31.2 million people in 2021.^2^ Recent estimates found that 30% of older adults living in underprivileged areas of Brazil experienced depressive symptoms, and approximately one-third of them had received a previous diagnosis.^3^

The COVID-19 pandemic exacerbated this scenario of mental health issues, increasing the burden of depression globally.^4^ For example, a meta-analysis conducted during the COVID-19 pandemic (2020-2021) estimated a prevalence of depression of 35% in Latin America.^5^ The pandemic also had a negative impact on access to healthcare. In Brazil, there was an acute drop in non-COVID services in the health system,^6^ partly due to the country’s unpreparedness to provide remote care.

The World Health Organization recommends primary care as the first step of depression management.^7^ Task-shared and collaborative care programmes led by trained non-mental health specialists have been shown to be cost-effective.^8–10^ However, these programmes require active and in-person participation of health professionals, which is not feasible with social distancing or when other priorities take precedence. In addition, the professionals providing routine primary care in LMIC are too busy to add and offer collaborative care to all older adults with depression.^11,12^

A useful strategy to improve access to treatment for depression in primary care without increasing health professionals’ workloads is the use of self-help programmes based on psychosocial techniques and delivered by digital technologies. Self-help programmes are low-cost alternatives with minimal or no contact with health professionals.^13,14^ However, there is still little evidence to support the effectiveness of self-help digital psychosocial interventions for older adults with depression in diverse primary care settings, especially in LMIC.^15^

We therefore developed an automated self-help programme (Viva Vida) for older adults with depression.^16^ Viva Vida was adapted from a task-shared, collaborative care psychosocial programme, named PROACTIVE, that we developed and investigated in Brazil.^17^ Thus, we conducted a randomised controlled trial (RCT) in primary care in Guarulhos, Brazil, to evaluate the effectiveness of the Viva Vida Programme, a six-week digital self-help psychosocial intervention delivered by automated WhatsApp messaging for the treatment of older adults with depression.^16^

## Methods

### Study design and participants

PRODIGITAL-D was a two-arm individually-randomised controlled trial with 1:1 allocation, conducted in the 24 largest primary care clinics, known as Unidades Básicas de Saúde (UBSs) in underprivileged areas of Guarulhos, Brazil. The city is part of the metropolitan region of Sao Paulo with a population of around 1.4 million people.

We recruited individuals who were 60+ years, registered with a participating UBS, able to receive and listen to WhatsApp messages, and who screened positive for depressive symptomatology (9-item Patient Health Questionnaire, PHQ-9 score≥10) with at least one core symptom of depression (low mood or anhedonia, i.e. a PHQ-2 score≥1).^18,19^ Exclusion criteria included significant visual or hearing impairments that would hinder the ability to comprehend messages on a mobile phone, acute suicidal risk identified during the screening assessment, another older adult living in the same household already participating in the RCT, and previous participation in the PROACTIVE trial. Verbal consent was obtained before the screening assessment and before the invitation to participate in the trial, both conducted by phone. The consent was audio-recorded after authorisation from the older adult. Details of the protocol are published elsewhere.^16^

This study was approved by the Ethics Committee of the Hospital das Clínicas da Faculdade de Medicina da Universidade de Sao Paulo – HCFMUSP (CAPPesq, ref: 4.097.596, first approved on 10 March 2021) and authorised by the Guarulhos Health Secretary.

### Randomisation and masking

Participants were stratified on age group (60-69 years, 70+ years), gender (male, female) and depression severity (PHQ-9 categories based on scores: 10-14, 15-19, 20+). The allocation sequence was generated using random permuted blocks with random block sizes by members of the research team not involved in the data collection (CAN and TJP). The ‘randomization’ module of the Research Electronic Data Capture (REDCap) was used to conceal the allocation sequence and randomise individuals after they consented to participate in the trial.

Research assistants involved in recruitment and follow-up data collection were masked to trial allocation. Two different groups of research assistants were responsible for either the first or the second follow-up assessment. Researchers responsible for scheduling and delivering messages were not masked to trial allocation but had no role in collecting inclusion or outcome data.

### Procedures

All assessments were conducted remotely. The names and phone numbers of all older adults registered with the 24 participating UBSs were provided by the Guarulhos Health Secretariat. After excluding duplicate entries and those with no phone numbers, individuals were assigned a random ID when entered into our system. First, research assistants sent a WhatsApp message to older adults according to the (randomly ordered) list of names to search for valid WhatsApp numbers. Secondly, those who received the pre-screening WhatsApp messages were approached by telephone to be screened for depressive symptomatology (using the PHQ-2).^18^ Individuals with a PHQ-2 score≥1 were asked the other seven items of the PHQ-9 questionnaire.^19^ Thirdly, those who scored PHQ-9≥10 completed a baseline assessment. Finally, those who were able to complete the baseline assessment were invited to participate in the trial. Invitation to participate could occur in the same call as the baseline assessment, or in an additional call no more than 28 days after the PHQ-9 screening. Follow-up data were collected by phone at three (weeks 12 to 16) and five months (weeks 20 to 24) after receiving the first message (intervention arm) or the single message (control arm).

### Interventions

Participants in both study arms continued receiving the usual primary care provided by the UBS. Health professionals had no role in the Viva Vida programme and the research team did not interfere with the care provided by the UBS.

### Viva Vida programme

Participants allocated to the intervention arm received an automated self-help psychosocial intervention delivered via WhatsApp. A total of 48 messages with psychoeducation and/or behavioural activation contents were sent across four days a week for six weeks, one in the morning and one in the afternoon. The contents about depressive symptoms and strategies to cope with them were introduced to participants by an audio message of around three minutes using storytelling techniques. Visual messages were used to review and reinforce the audio message contents. At the end of each week, participants received an extra message with a simple question about their opinion and experience on the programme, and they were able to provide feedback using the WhatsApp quick reply tool. This tool included up to three pre-defined answer options. Additionally, participants received one message with the technical support contact in case they experienced technical issues. A <u>video excerpted from a national TV show</u> presenting details on the programme is available online.

### Enhanced Usual Care

The control arm participants were sent a single psychoeducational message. The audio message provided information about depression and simple ways to deal with depressive symptoms. It also offered additional tips encouraging participants to live a healthier life, such as improving sleep and maintaining a balanced diet, and it advised participants to seek healthcare if they felt they needed additional support.

### Data collection and management

Depressive symptomatology was assessed with the PHQ-9,^19^ anxiety symptomatology with the Generalized Anxiety Disorder-7 (GAD-7),^20^ loneliness with the 3-item University of California, Los Angeles (UCLA) loneliness scale (3-item UCLA),^21^ quality of life with the European Quality of Life five-dimensional questionnaire, five-level version (EQ-5D-5L),^22^ and capability wellbeing with the ICEpop CAPability measure for Older people (ICECAP-O).^23^. No value set of the EQ-5D-5L and ICECAP-O measures is defined for the Brazilian population; thus, we used the tariff values for the Uruguayan population^24^ and the UK population,^25^ respectively. All these instruments were applied at baseline, three– and five-month follow-up assessments. Sociodemographic and clinical characteristics were also collected at baseline. Information on medical appointments and hospitalisations were collected at each follow-up assessment. Severe adverse events were assessed for all participants at three and five months.

Data were collected and managed using REDCap^26,27^ hosted at the Hospital das Clínicas da Faculdade de Medicina da Universidade de Sao Paulo. REDCap is a secure, web-based software platform designed to support data capture for research studies. A system was developed to collect and store data from the WhatsApp messages.

### Outcomes

Recovery from depression (PHQ-9 score<10) at three months was defined as the primary outcome. Secondary outcomes included recovery from depression at five months, reduction in PHQ-9 scores by at least 50% between baseline and follow-up visits at both three and five months. We also evaluated effects of the intervention on the continuous scores of anxiety symptomatology (GAD-7), loneliness (3-item UCLA), health-related quality of life (EQ-5D-5L), and capability wellbeing (ICECAP-O) at three and five months.

### Sample size

With an assumed 15% attrition, 440-500 randomised individuals would yield 80-85% power to detect a 15-percentage point difference in depression recovery rates between the control and intervention arms at three months (25% versus 40%)^9,28^ using a two-sided 5% significance level.

### Statistical methods

Descriptive statistics were obtained using complete case. All comparative analyses were performed using intention-to-treat principles with imputed data in regression models, aligned to the Statistical Analysis Plan (SAP).^29^ Logistic regression was used for all binary outcomes (including the primary); linear regression was used for continuous outcomes. All regression analyses were adjusted for stratification (above or below the median proportion of adults aged 70 years or older, gender, and depression severity at baseline (PHQ-9 categories)). All analyses that had an outcome other than the PHQ-9 were also adjusted for the baseline score of the corresponding outcome.

Pre-specified subgroup analyses at both follow-up assessments were investigated using likelihood ratio tests for interactions between randomised arm and the following variables: gender; age; educational level; comorbid physical illness (hypertension, diabetes, or both); and baseline PHQ-9 severity categories. We also ran a model for the primary outcome (recovery at three months) adjusting for elapsed time between consenting into the trial and the first follow-up assessment.

Missing data were replaced using multiple imputation by chained equations (MICE) under the assumption that data were missing at random. MICE models were used for all primary and secondary outcomes. Supplemental material provides details of the methods used for the missing data analyses including sensitivity analyses as well as results comparing estimates using complete cases versus imputed data (eTables 1-8).

Complier Average Case Effect (CACE) analysis^30^ using an instrumental variable estimator and imputed data was applied to estimate the effect on depression outcomes of the number of messages electronically recorded as being ‘opened’. The SAP pre-specified the ‘minimum therapeutic dose’ as “listening to more than half of the messages received”.^29^ In the event, the available data related to how many of the 48 psychosocial messages were electronically recorded as being ‘opened’. We therefore operationalised the SAP threshold as participants who ‘opened’ 36 or more messages (75% of the programme) versus ‘opening’ 35 or fewer messages. The CACE analyses were conducted using PHQ-9 score at three and five months, adjusting for stratification. We also conducted sensitivity analyses using thresholds of ‘opening’ at least half of the messages (24+) versus 23 messages or fewer, and ‘opening’ all 48 messages versus not doing so.

Regression diagnostics were run for both the logistic regression and the linear regression models. The Box-Tidwell test was run after the logistic regression models to test whether the logit transform is a linear function of the predictors for the different models. Normality assumptions for all linear regression models were evaluated by examining the residual plots. Statistical tests were two-sided, and all analyses were conducted using Stata 17 (StataCorp).

## Results

Data collection took place between 8 September 2021 and 26 September 2022. A list with 36,278 older adults registered with the 24 primary care clinics was provided by the Guarulhos Health Secretariat. Among the 13,968 (38.5%) individuals with a valid WhatsApp number and screened for eligibility by phone: 2,720 (19.5%) did not meet eligibility criteria (of whom 1,717 did not have depressive symptomatology); 562 (4.0%) declined participation; and 10,083 (72.2%) could not be contacted (Figure 1). A total of 603 participants were recruited, 298 randomised to the intervention arm and 305 to the control arm. With variations driven largely by differences in catchment populations, between five and 45 participants were recruited in each UBS, though the majority of them recruited 20-30 participants.

**Figure 1.**
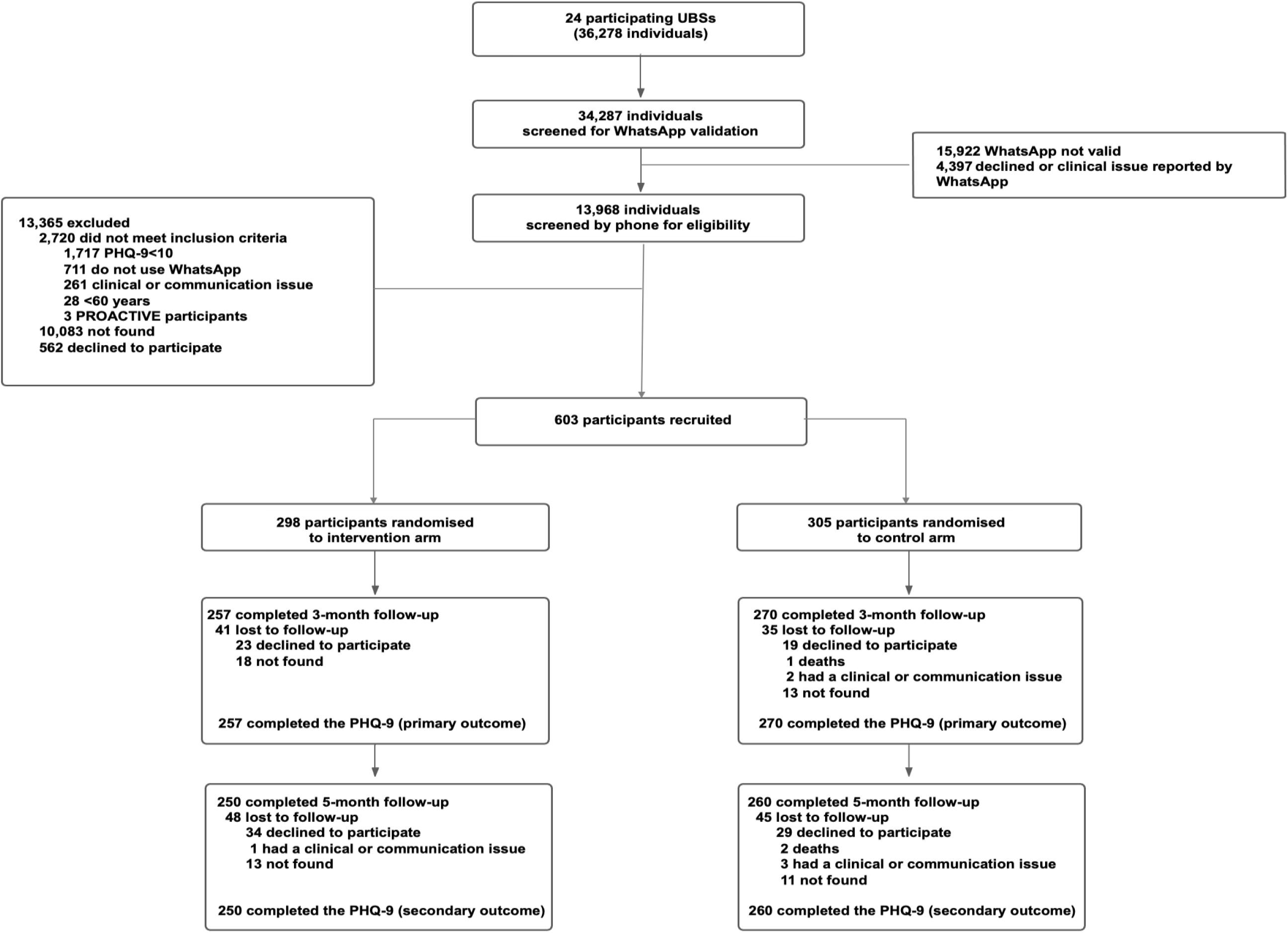
Participant flow through the trial.

A total of 527 (87.4%) participants recruited were followed-up at three months and the primary outcome was available for all of them, with 41 (13.8%) lost to follow-up in the intervention group and 35 (11.5%) in the control group. At the five-month follow-up, 510 (84.6%) participants were assessed.

Descriptive statistics of sociodemographic and other relevant characteristics suggest there were no major differences between trial arms at baseline (Table 1); eTable 9 and eTable10 demonstrates that baseline characteristics remained balanced at both follow-up visits. The majority of participants were 60-69 year-old women, of whom the majority had less than eight years of education and were earning minimum wage. The proportion of participants with self-reported hypertension was high (69.5% and 72.8% in the intervention and control arms respectively) and to a lesser degree diabetes (41.3% and 39.7% respectively).

**Table 1.** Baseline characteristics in the intervention and control arms.

| Number/total sample size (%) |  |  |
| --- | --- | --- |
|  | Viva Vida (n=298) | Enhanced Usual Care (n=305) |
| **Female** | 225/298 (75.5) | 226/305 (74.1) |
| **Age group** |  |  |
| 60-69 years | 251/298 (84.2) | 251/305 (82.3) |
| 70+ years | 47/298 (15.8) | 54/305 (17.7) |
| **Education** |  |  |
| None | 45/297 (15.2) | 51/302 (16.9) |
| 1-4 years | 83/297 (27.9) | 84/302 (27.8) |
| 5-8 years | 94/297 (31.6) | 85/302 (28.1) |
| >8 years | 75/297 (25.3) | 82/302 (27.2) |
| **Personal income** |  |  |
| Up to 1 MW | 197/298 (66.1) | 191/305 (62.6) |
| >1-2 MW | 60/298 (20.1) | 56/305 (18.4) |
| >2 MW | 41/298 (13.8) | 58/305 (19.0) |
| **Hypertension (self-reported)** | 207/298 (69.5) | 222/305 (72.8) |
| **Diabetes (self-reported)** | 123/298 (41.3) | 121/305 (39.7) |
| **Receiving pharmacological treatment for depression (self-reported)** | 38/298 (12.8) | 46/305 (15.1) |
| **PHQ-9 severity** |  |  |
| 10-14 | 120/298 (40.3) | 119/305 (39.0) |
| 15-19 | 99/298 (33.2) | 100/305 (32.8) |
| 20+ | 79/298 (26.5) | 86/305 (28.2) |

*Abbreviations:* MW: minimum wage (in 2021, the minimum wage in Brazil was BRL1110 (approximately US\$213); PHQ-9: 9-item Patient Health Questionnaire for depression.

Data collected during the follow-up visits showed that six participants (2.0%) in each arm reported suicidal ideation. Eighteen participants (6.0%) in the intervention arm and 20 participants (6.6%) in the control arm were hospitalised for any reason during the trial. Also, two deaths (0.7%) were reported in the control arm. All events were considered not related to either the intervention or study participation.

### Primary outcome

The results from the primary analysis demonstrated a clear beneficial intervention effect in recovering from depression (PHQ-9 score<10) at three months, with 109 (42.4%) in the intervention arm recovering from depression compared with 87 (32.2%) in the control arm (Table 2). The adjusted odds ratio for this difference after imputing missing values was 1.56 (95% confidence interval (CI): 1.07, 2.27; *P*=.021). Complete case analyses led to very similar findings (eTable5).

**Table 2.**
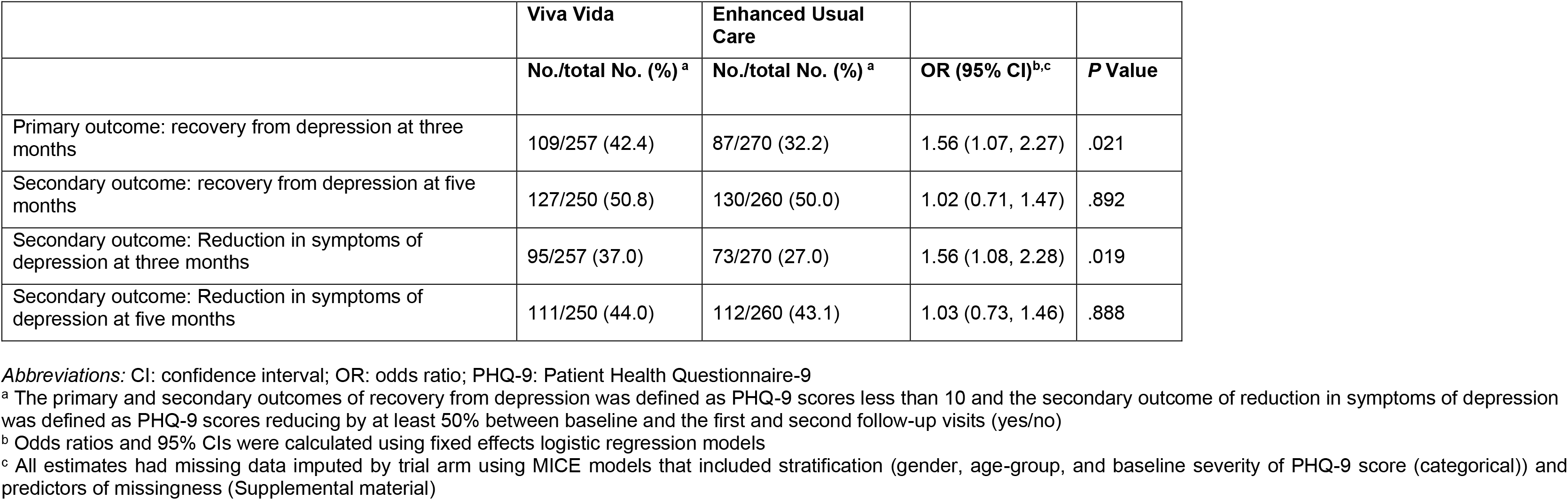
Primary and secondary depression outcomes at three– and five-month follow-up visits of recovery from depression and of improvement in symptoms of depression (PHQ-9 scores reduced by at least 50% between baseline and first and second follow-up visits)

### Secondary outcomes

From Table 2, the proportions recovering from depression at five months were virtually identical in the two groups, at 50.8% and 50.0% for intervention and control arms, respectively (adjusted odds ratio: 1.02; 95% CI: 0.71, 1.47; *P*=.892). Table 2 also shows that the secondary outcome of PHQ-9 scores improving by at least 50% demonstrated a benefit of the intervention at three months (1.56; 95% CI: 1.08, 2.28; *P*=.019), but not at five months (1.03; 95% CI: 0.73, 1.46; *P*=.888). Complete case analyses showed very similar results (eTable 5 and eTable 6). There was no evidence of differences for any other secondary outcomes at the three-month or five-month follow-up visits, either using imputed data (Table 3) or in the complete case analyses (eTable 7 and eTable 8).

**Table 3.** Secondary outcomes including depressive symptomatology (PHQ-9), anxiety symptomatology (GAD-7), health-related quality of life (EQ-5D-5L), and capability wellbeing in older adults (ICECAP-O), and perceived loneliness (UCLA), at three– and five-month follow-up.

| Outcome | Baseline |  | Three-month follow-up |  |  |  | Five-month follow-up |  |  |  |
| --- | --- | --- | --- | --- | --- | --- | --- | --- | --- | --- |
|  | Viva Vida | Enhanced Usual Care | Viva Vida | Enhanced Usual Care | Coefficient (95% CI)<sup>a, b</sup> | P Value | Viva Vida | Enhanced Usual Care | Coefficient (95% CI)<sup>a, b</sup> | P Value |
| **PHQ-9, Number** | 298 | 305 | 257 | 270 | -1.14<br>(-2.34, 0.05) | .060 | 250 | 260 | -0.22<br>(-1.37, 0.93) | .703 |
| Mean (SD) | 16.39<br>(4.66) | 16.68<br>(4.83) | 11.72<br>(7.72) | 12.92<br>(7.13) |  |  | 10.01<br>(7.07) | 10.41<br>(7.01) |  |  |
| **GAD-7, Number** | 285 | 283 | 257 | 247 | -0.48<br>(-1.75, 0.77) | .450 | 250 | 260 | -0.48<br>(-1.75, 0.77) | .450 |
| Mean (SD) | 14.46<br>(4.62) | 14.36<br>(4.75) | 10.72<br>(9.33) | 11.07<br>(6.43) |  |  | 9.40<br>(6.79) | 9.49<br>(6.66) |  |  |
| **EQ-5D-5L, Number** | 296 | 305 | 256 | 266 | 0.024<br>(-0.020, 0.067) | .091 | 247 | 259 | 0.026<br>(-0.005, 0.077) | .085 |
| Mean (SD) | 0.789<br>(0.182) | 0.794<br>(0.170) | 0.812<br>(0.189) | 0.801<br>(0.171) |  |  | 0.840<br>(0.164) | 0.825<br>(0.171) |  |  |
| **ICECAP-O, Number** | 295 | 300 | 254 | 266 | 0.018<br>(-0.009, 0.045) | .404 | 246 | 259 | 0.002<br>(-0.024, 0.029) | .859 |
| Mean (SD) | 0.614<br>(0.172) | 0.621<br>(0.190) | 0.655<br>(0.174) | 0.642<br>(0.194) |  |  | 0.665<br>(0.165) | 0.667<br>(0.172) |  |  |
| **UCLA, Number** | 298 | 305 | 256 | 267 | 0.52<br>(-0.28, 1.33) | .199 | 250 | 259 | 0.22<br>(-0.07, 0.51) | .138 |
| Mean (SD) | 6.04<br>(1.91) | 6.16<br>(1.92) | 6.05<br>(6.33) | 5.34<br>(1.94) |  |  | 5.43<br>(1.97) | 5.25<br>(1.97) |  |  |

Abbreviations: EQ-5D-5L, 5-level EuroQol health-related quality of life questionnaire; GAD-7, 7-item General Anxiety Disorder questionnaire; ICECAP-O, ICEpop CAPability measure for older people; PHQ-9, Patient Health Questionnaire-9; SD: standard deviation.
PHQ-9 scores range from 0 to 27, with higher scores representing more severe symptoms of depression
GAD-7 scores range from 0 to 21 with higher scores representing more severe symptoms of anxiety
EQ-5D-5L scores range from -0.264 to 1, with higher scores representing higher quality of life
ICECAP-O scores range from 0 to 1, with higher scores representing greater levels of capability wellbeing
<sup>a</sup> Difference in means were estimated using linear regression models, adjusted for the baseline assessment of the corresponding outcome and the stratified variable (gender, age-group, and baseline severity of PHQ-9 score (categorical))
<sup>b</sup> All estimates had missing data imputed separately, by trial arm, using MICE models that included predictors of missingness (Supplemental material), stratification (gender, age-group, and baseline severity of PHQ-9 score (categorical)), and any imbalances in outcome measure at baseline

### Additional analyses

The electronic system capturing the number of messages that were ‘opened’ indicated that 226 (75.8%) participants in the intervention arm ‘opened’ most of the messages (36 or more out of 48), 237 (79.5%) ‘opened’ half of them or more, and 173 (58.1%) ‘opened’ them all. Although the confidence interval at three months only just includes the null, the main CACE analysis (evaluating the effect of ‘opening’ most of the messages), did not yield clear evidence of an improvement in mean PHQ-9 scores for these participants compared with others (Table 4) at either the three– or five-month follow-ups. The sensitivity analyses led to the same conclusion as the main CACE analysis (Table 4). There was no evidence of differential effects according to the pre-specified subgroup analyses (eTable 11).

**Table 4.**
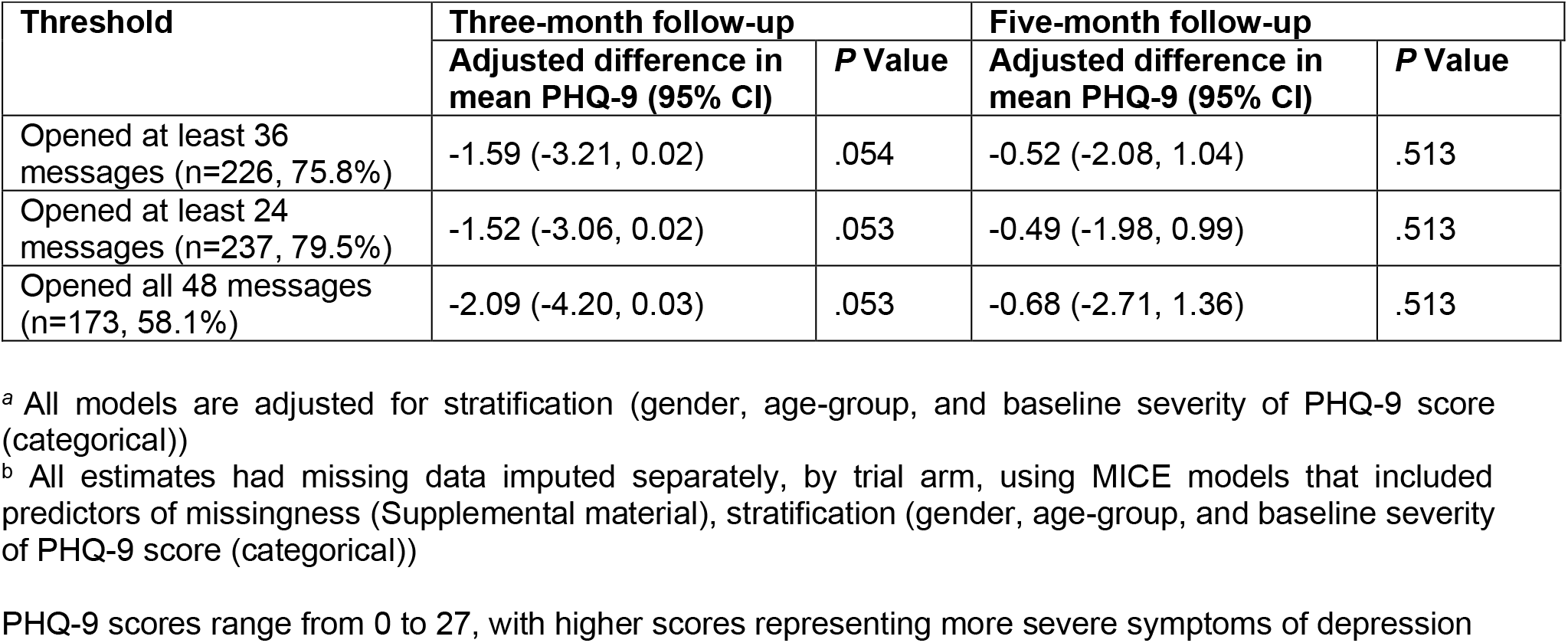
Complier-average causal effects (CACE) analyses^a,b^ on the mean PHQ-9 scores at three– and five-month follow-ups using electronic recordings.

| Threshold | Three-month follow-up |  | Five-month follow-up |  |
| --- | --- | --- | --- | --- |
|  | Adjusted difference in mean PHQ-9 (95% CI) | P Value | Adjusted difference in mean PHQ-9 (95% CI) | P Value |
| Opened at least 36 messages (n=226, 75.8%) | -1.59 (-3.21, 0.02) | .054 | -0.52 (-2.08, 1.04) | .513 |
| Opened at least 24 messages (n=237, 79.5%) | -1.52 (-3.06, 0.02) | .053 | -0.49 (-1.98, 0.99) | .513 |
| Opened all 48 messages (n=173, 58.1%) | -2.09 (-4.20, 0.03) | .053 | -0.68 (-2.71, 1.36) | .513 |

<sup>a</sup> All models are adjusted for stratification (gender, age-group, and baseline severity of PHQ-9 score (categorical))
<sup>b</sup> All estimates had missing data imputed separately, by trial arm, using MICE models that included predictors of missingness (Supplemental material), stratification (gender, age-group, and baseline severity of PHQ-9 score (categorical))
PHQ-9 scores range from 0 to 27, with higher scores representing more severe symptoms of depression

## Discussion

To the best of our knowledge, this is the first RCT assessing a digital psychosocial intervention delivered by WhatsApp to older adults in a LMIC. The six-week Viva Vida programme showed increased odds of recovery from depression among older adults at the (primary) three-month follow-up compared with a single psychoeducation message. The digital intervention also showed improvement on a reduction of at least 50% in PHQ-9 scores at this follow-up. No differences between the two arms on these outcomes were observed at five months. Moreover, levels of anxiety symptomatology (GAD-7), loneliness (3-item UCLA), health-related quality of life (EQ-5D-5L) and capability wellbeing (ICECAP-O) were similar in the two arms at both the three– and five-month follow-ups.

The use of digital strategies to deliver a behaviour activation intervention has demonstrated small to medium effects on depression, anxiety and quality of life in adults.^31^ Similar to our findings, one study conducted in middle-to-older adults with depression and comorbid hypertension or diabetes in Brazil and Peru showed that a six-week smartphone-based intervention guided by nurse assistants improved depressive symptoms at three, but not at six, months compared with enhanced usual care.^32^ Both studies suggest that digital interventions based on behavioural activation accelerate the recovery from depression in the short-term among middle-to-older adults.

WhatsApp was the tool selected to deliver our intervention as it is widely used by the target population. The use of a well-known and accepted tool enables the receipt of messages without requiring the download of a new application or the acquisition of additional digital skills. Therefore, older adults with relatively low digital literacy level or without access to high-speed internet connection can also benefit from this intervention. Our choice for the popular messaging application is consistent with findings from a recent systematic review on digital interventions for mental health in older adults suggesting that successful interventions use simple technologies.^15^ However, despite potentially improving adherence to and engagement with the intervention,^33^ this popular application imposes some limitations for collecting data for the study. For instance, although it is possible to know what proportion of the messages were ‘opened’, it is not possible to collect data on the extent to which the messages were listened to, let alone the actual adherence to the intervention messages, as WhatsApp only provides data on messages ‘opened’.

Human support has also been reported as a significant component of effective digital mental health interventions.^15^ However, the Viva Vida programme did not include such support to deliver the self-help intervention, as it was intended to be a low-intensity and low-budget intervention that offered a first step for the management of depression in primary care. We found improved outcomes of depression at three months only, but with an effect size similar to interventions supported by nurse assistants^32^ or lay counsellors.^34^ Although the point estimate of the intervention effect (10.2 percentage points in absolute terms) was lower than that specified as the original target difference (15 percentage points), we nonetheless contend that this is an important finding due to the high potential for scalability of this programme. In LMIC settings where access to mental health care is limited, self-help programmes can help filling the treatment gap when there are no other available treatment options. We do not expect it to be suitable for everyone, however, and those who do not benefit from this type of intervention would then need to be referred to a more intense programme, such as the PROACTIVE programme,^8^ in which CHWs visits may have played an important role in prolonging the benefits of the intervention after its conclusion.

This study has some other limitations. Individuals with a lower digital literacy and/or without access to WhatsApp, individuals with cognitive impairment and those with vision or hearing issues were not included in this study due to the design of the self-help digital intervention. Major adaptations are needed in order to deliver a more appropriate intervention to reach such individuals.

## Conclusions

Our RCT provides evidence on the effectiveness of a simple and short self-help digital intervention for depression in older adults that does not require the active support of health professionals. An additional 10 percentage point recovery rate with a low-cost intervention such as Viva Vida can lead to a major impact if and when this is implemented at larger scale in other settings in Brazil or other LMICs, reaching individuals with depressive symptoms who do not have access to mental health care. Further analyses are investigating the cost-effectiveness and the implementation outcomes of the Viva Vida programme to understand its full potential benefits and costs.

## Data Availability

All data produced in the present study are available upon reasonable request to the authors

https://www.youtube.com/watch?v=oW5LWSfZ3dQ&t=19s

## Appendices

### Sensitivity analyses – missing data analyses

#### Methods

Initially, patterns of missingness were investigated by comparing missing with complete data for the baseline demographic characteristics and morbidity questionnaires with the outcome of depression, by trial arm, at both follow-up visits. In order to reduce bias and loss of information, we used multiple imputation by chained equations (MICE) with 50 imputations, as implemented in the MI command in Stata under the assumption that data were missing at random (MAR).^1^ Variables used in the MICE models consisted of the outcome recovered from depression, covariates described in the Statistical Analysis Plan (age group, severity of PHQ-9, gender, treatment arm) and variables found to be predictors of missingness.^2,3^ Predictors of missingness include the following: hypertension at baseline, income, and levels of enjoyable activities participants engaged in. Baseline scores for corresponding outcome models including depressive symptomatology (PHQ-9), anxiety symptomatology (GAD-7), health-related quality of life (EQ-5D-5L), capability wellbeing (ICECAP-O), and levels of loneliness captured with 3-item UCLA scores were also included.

To assess the sensitivity of our findings against modest departures from the MAR assumption, a weighted sensitivity analysis using the Selection Model Approach was applied.^4–6^ Briefly, once data had been imputed under MAR, parameter estimates from each imputed dataset were reweighted to allow for the data to be missing not at random (MNAR). To test the stability of our model, we considered different degrees of departure from the MAR assumption by considering plausible values of δ ranging from 0.10 to 0.40. This range corresponds to odds ratios for the data being observed when a participant recovered from depression compared to when it did not, ranging from 1.11 to 1.50 (i.e. exponential of 0.10 and 0.40).

#### Results

##### General

eTables 1 and 2 demonstrate findings from the analysis comparing baseline demographics and baseline measures for the secondary outcomes between participants with complete data, and those with missing data, at three– and five-months follow-up visits respectively.

**eTable 1:**
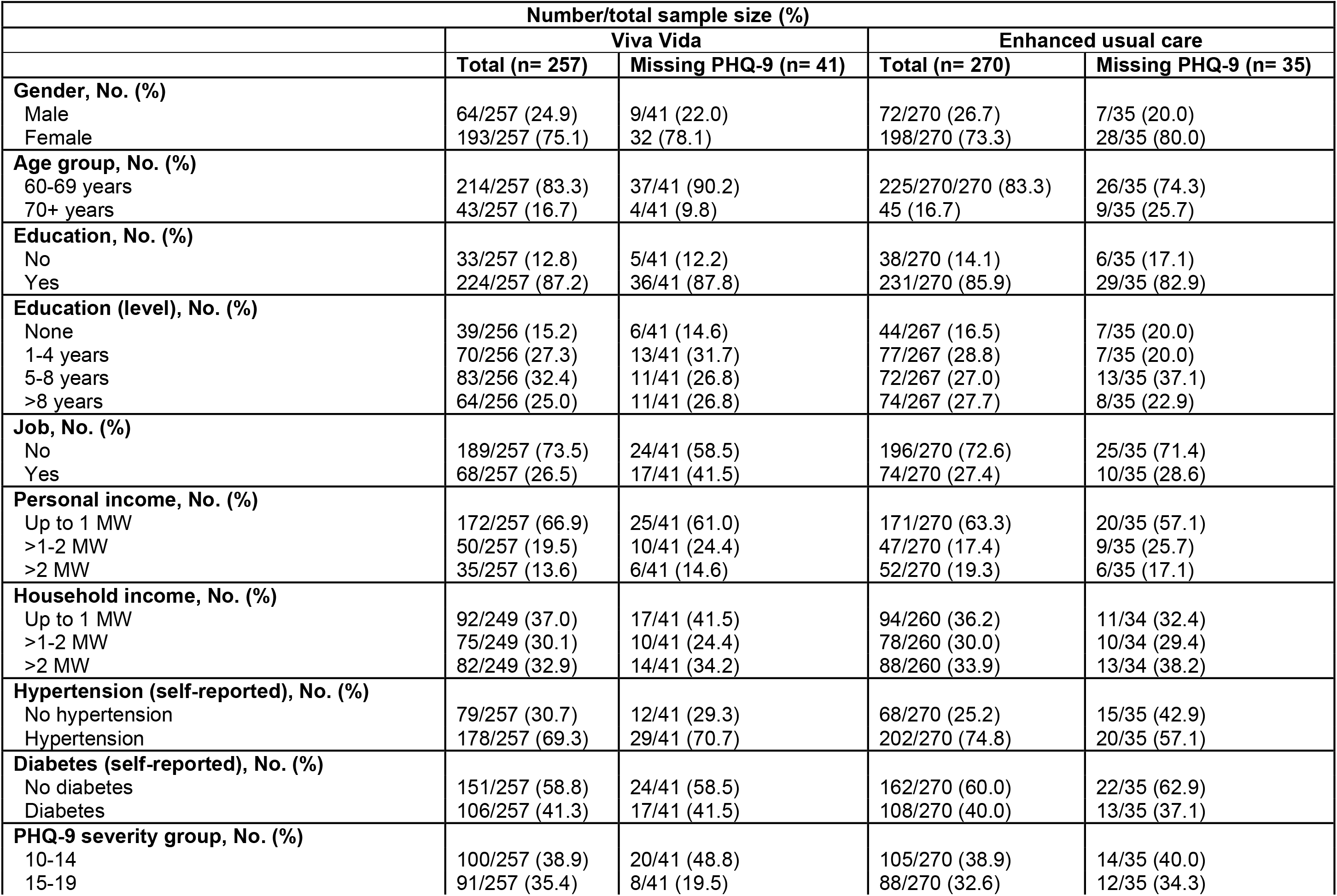
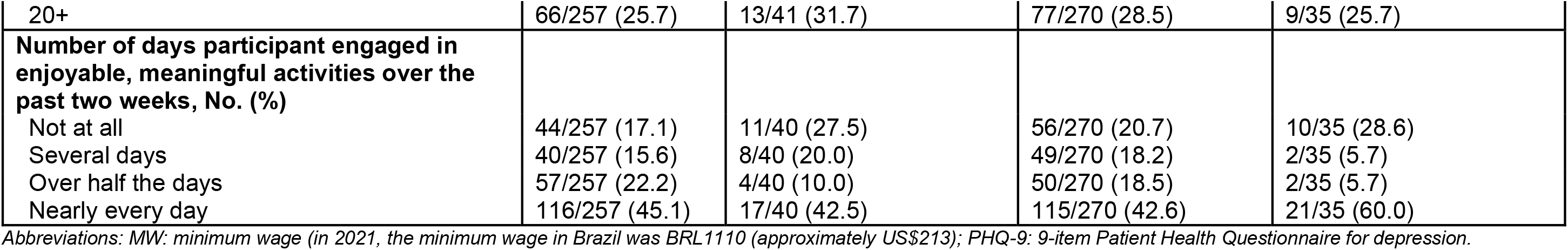
Comparison between participants with complete data and participants with missing data at the three-month follow-up visit.

**eTable 2:** Comparison between participants with complete data and participants with missing data at the five-month follow-up visit.

| Number/total sample size (%) |  |  |  |  |
| --- | --- | --- | --- | --- |
|  | Viva Vida |  | Enhanced usual care |  |
|  | Total (n= 250) | Missing PHQ-9 (n= 48) | Total (n= 260) | Missing PHQ-9 (n= 45) |
| **Gender, No. (%)** |  |  |  |  |
| Male | 64/250 (25.6) | 9/48 (18.8) | 72/260 (27.7) | 7/45 (15.6) |
| Female | 186/250 (74.4) | 39/48 (81.3) | 188/260 (72.3) | 38/45 (84.4) |
| **Age group, No. (%)** |  |  |  |  |
| 60-69 years | 210/250 (84.0) | 41/48 (85.4) | 216/260 (83.1) | 35/45 (77.8) |
| 70+ years | 40/250 (16.0) | 7/48 (14.6) | 44/260 (16.9) | 10/45 (22.2) |
| **Education, No. (%)** |  |  |  |  |
| No | 32/250 (12.8) | 6/48 (12.5) | 36/260 (13.9) | 8/45 (17.8) |
| Yes | 218/250 (87.2) | 42/48 (87.5) | 223/260 (86.1) | 37/45 (82.2) |
| **Education (level), No. (%)** |  |  |  |  |
| None | 38/249 (15.3) | 7/48 (14.6) | 41/257 (16.0) | 10/45 (22.2) |
| 1-4 years | 63/249 (25.3) | 20/48 (41.7) | 75/257 (29.2) | 9/45 (20.0) |
| 5-8 years | 81/249 (32.5) | 13/48 (27.1) | 70/257 (27.2) | 15/45 (33.3) |
| >8 years | 67/249 (26.9) | 8/48 (16.7) | 71/257 (27.6) | 11/45 (24.4) |
| **Job, No. (%)** |  |  |  |  |
| No | 184/250 (73.6) | 29/48 (60.4) | 191/260 (73.5) | 30/45 (66.7) |
| Yes | 66/250 (26.4) | 19/48 (39.6) | 69/260 (26.5) | 15/45 (33.3) |
| **Personal income, No. (%)** |  |  |  |  |
| Up to 1 MW | 163/250 (65.2) | 34/48 (70.8) | 162/260 (62.3) | 29/45 (64.4) |
| >1-2 MW | 50/250 (20.0) | 10/48 (20.8) | 46/260 (17.7) | 10/45 (22.2) |
| >2 MW | 37/250 (14.8) | 4/48 (8.3) | 52/260 (20.0) | 6/45 (13.3) |
| **Household income, No. (%)** |  |  |  |  |
| Up to 1 MW | 89/242 (36.8) | 20/48 (41.7) | 90/250 (36.0) | 15/44 (34.1) |
| >1-2 MW | 70/242 (28.9) | 15/48 (31.3) | 77/250 (30.8) | 11/44 (25.0) |
| >2 MW | 83/242 (34.3) | 13/48 (27.1) | 83/250 (33.2) | 18/44 (40.9) |
| **Hypertension (self-reported), No. (%)** |  |  |  |  |
| No hypertension | 82/250 (32.8) | 9/48 (18.8) | 67/260 (25.8) | 16/45 (35.6) |
| Hypertension | 168/250 (67.2) | 39/48 (81.3) | 193/260 (74.2) | 29/45 (64.4) |
| **Diabetes (self-reported), No. (%)** |  |  |  |  |
| No diabetes | 144/250 (57.6) | 31/48 (64.6) | 158/260 (60.8) | 26/45 (57.8) |
| Diabetes | 106/250 (42.4) | 17/48 (35.4) | 102/260 (39.2) | 19/45 (42.2) |
| **PHQ-9 severity group, No. (%)** |  |  |  |  |
| 10-14 | 102/250 (40.8) | 18/48 (37.5) | 104/260 (40.0) | 15/45 (33.3) |
| 15-19 | 87/250 (34.8) | 12/48 (25.0) | 87/260 (33.5) | 13/45 (28.9) |
| 20+ | 61/250 (24.4) | 18/48 (37.5) | 69/260 (26.5) | 17/45 (37.8) |
| **Number of days participant engaged in enjoyable, meaningful activities over the past two weeks, No. (%)** |  |  |  |  |
| Not at all | 42/249 (16.9) | 13/48 (27.1) | 56/260 (21.5) | 10/45 (22.2) |
| Several days | 41/249 (16.5) | 7/48 (14.6) | 49/260 (18.9) | 2/45 (4.4) |
| Over half the days | 54/249 (21.7) | 7/48 (14.6) | 43/260 (16.5) | 9/45 (20.0) |
| Nearly every day | 112/249 (45.0) | 21/48 (43.8) | 112/260 (43.1) | 24/45 (53.3) |

Abbreviations: MW: minimum wage (in 2021, the minimum wage in Brazil was BRL1110 (approximately US\$213); PHQ-9: 9-item Patient Health Questionnaire for depression.

##### Sensitivity analysis checking the missing at random (MAR) assumption for the primary outcome

Results from the sensitivity analysis testing the MAR assumption for the primary outcome of recovery from depression at three months suggest that estimates moved away from the null. However, at five months, the opposite occurred, suggesting a different mechanism for missing data (participants who recovered from depression, more likely to be missing at follow-up, compared to participants who did not recover from depression).

**eTable 3:**
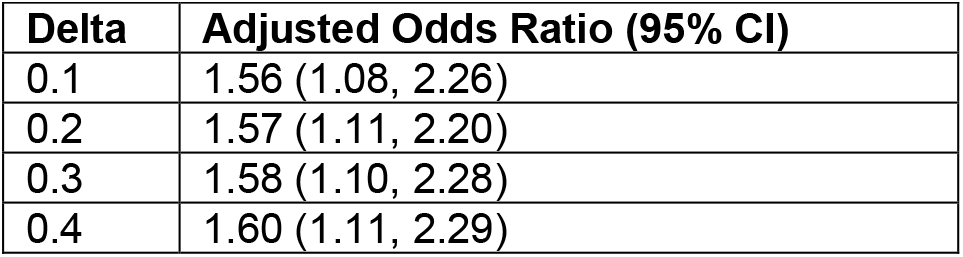
Model 1 – Adjusted odds ratios (95% CI) for different departures from the missing at random assumption, for the outcome variable of recovery from depression at three months assuming greater probability of the outcome being missing when depression was present.

| Delta | Adjusted Odds Ratio (95% CI) |
| --- | --- |
| 0.1 | 1.56 (1.08, 2.26) |
| 0.2 | 1.57 (1.11, 2.20) |
| 0.3 | 1.58 (1.10, 2.28) |
| 0.4 | 1.60 (1.11, 2.29) |

**eTable 4:**
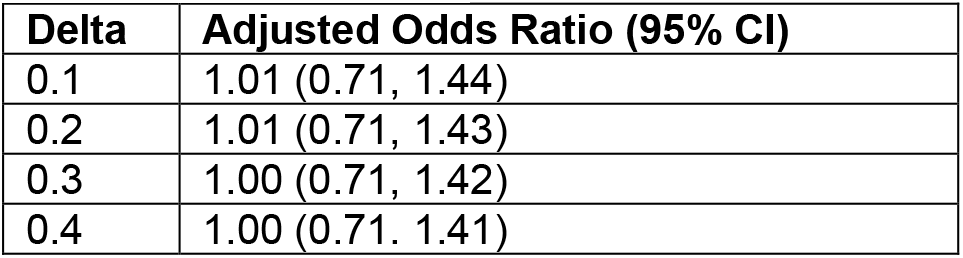
Model 2 – Adjusted odds ratios (95% CI) for different departures from the missing at random assumption, for the outcome variable of recovery from depression at five months assuming greater probability of the outcome being missing when depression was present.

| Delta | Adjusted Odds Ratio (95% CI) |
| --- | --- |
| 0.1 | 1.01 (0.71, 1.44) |
| 0.2 | 1.01 (0.71, 1.43) |
| 0.3 | 1.00 (0.71, 1.42) |
| 0.4 | 1.00 (0.71, 1.41) |

##### Results comparing estimates from MICE models with complete case analyses for both primary and secondary outcomes

**eTable 5:** Comparison of adjusted estimates from multiple imputed data using MICE models, with estimates from complete case analysis for the recovery of depression at three months (primary outcome) and five months (secondary outcome)

|  | Complete case analysis<sup>a,b,c</sup> |  | Imputed data from MICE analyses<sup>a,b,c</sup> |  |
| --- | --- | --- | --- | --- |
| Model | Odds ratio (95% CI) | P Value | Odds ratio (95% CI) | P Value |
| Recovery from depression at three months<sup>a,b</sup> (n=527) | 1.58 (1.09, 2.29) | 0.016 | 1.56 (1.07, 2.27) | 0.021 |
| Recovery from depression at five months<sup>a,b</sup> (n=510) | 1.02 (0.71, 1.46) | 0.911 | 1.02 (0.71, 1.47) | 0.892 |

<sup>a</sup> The primary and secondary outcome of recovery from depression was defined as PHQ-9 scores less than 10
<sup>b</sup> Odds ratios and 95% CIs were calculated using logistic regression models adjusted for gender, age-group, and baseline severity of PHQ-9 (categorical).
<sup>c</sup> All estimates had missing data imputed by trial arm using MICE models that included gender, age-group, baseline severity of PHQ-9 (categorical), and predictors of missingness

**eTable 6:** Comparison of adjusted estimates from multiple imputed data using MICE models, with estimates from complete case analysis for a reduction in PHQ-9 scores by at least 50% between baseline and three and five months (secondary outcomes)

|  | Complete case analysis<sup>a,b,c</sup> |  | Imputed data from MICE analyses<sup>a,b,c</sup> |  |
| --- | --- | --- | --- | --- |
| Model | Odds ratio (95% CI) | P Value | Odds ratio (95% CI) | P Value |
| Reduction in PHQ-9 scores by at least 50% between baseline and three months<sup>a,b</sup> (n=527) | 1.59 (1.10, 2.31) | 0.015 | 1.56 (1.08, 2.28) | 0.019 |
| Reduction in PHQ-9 scores by at least 50% between baseline and five months<sup>a,b</sup> (n=510) | 1.06 (0.75, 1.51) | 0.740 | 1.03 (0.73, 1.46) | 0.888 |

<sup>a</sup> The secondary outcome of reduction in symptoms of depression was defined as PHQ-9 scores reducing by at least 50% between baseline and the first and second follow-up visits (yes/no)
<sup>b</sup> Odds ratios and 95% CIs were calculated using logistic regression models adjusted for gender, age-group, and baseline severity of PHQ-9 (categorical).
<sup>c</sup> All estimates had missing data imputed by trial arm using MICE models that included gender, age-group, baseline severity of PHQ-9 (categorical), and predictors of missingness

**eTable 7:** Comparison of estimates from multiple imputed data using MICE models, with estimates from complete case analysis for secondary outcomes at three months.

| Model | Complete case analysis<sup>a,b</sup> |  | Imputed data from MICE analyses<sup>a,b</sup> |  |
| --- | --- | --- | --- | --- |
|  | Difference in means (95% CI) | P Value | Difference in means (95% CI) | P Value |
| PHQ-9 scores<sup>a</sup> | -1.09 (-2.28, 0.09) | 0.070 | -1.14 (-2.34, 0.05) | 0.060 |
| GAD-7 scores<sup>a</sup> | -0.37 (-1.67, 0.914) | 0.567 | -0.48 (-1.75, 0.77) | 0.450 |
| EQ-5D-5L scores<sup>a</sup> | 0.015 (-0.010, 0.039) | 0.246 | 0.024 (-0.020, 0.067) | 0.091 |
| ICECAP-O scores<sup>a</sup> | 0.021 (-0.006, 0.048) | 0.132 | 0.018 (-0.009, 0.045) | 0.404 |
| 3-item UCLA scores<sup>a</sup> | 0.54 (-0.20, 1.33) | 0.150 | 0.52 (-0.28, 1.33) | 0.199 |

<sup>a</sup> Differences in means were estimated using linear regression models, adjusted for relevant baseline assessment of corresponding outcome, gender, age-group, baseline severity of PHQ-9 (categorical)
<sup>b</sup> All estimates had missing data imputed separately, by trial arm, using MICE models that included predictors of missingness, gender, age-group, baseline severity of PHQ-9 score (categorical)
PHQ-9 scores range from 0 to 27, with higher scores representing more severe symptoms of depression
GAD-7 scores range from 0 to 21 with higher scores representing more severe symptoms of anxiety
EQ-5D-5L scores range from -0.264 to 1, with higher scores representing higher quality of life
ICECAP-O scores range from 0 to 1, with higher scores representing greater levels of capability wellbeing
3-item UCLA scores range from 3 to 9, with higher scores representing more severe levels of loneliness

**eTable 8:** Comparison of estimates from multiple imputed data using MICE models, with estimates from complete case analysis for secondary outcomes at five months.

| Model | Complete case analysis<sup>a,b</sup> |  | Imputed data from MICE analyses<sup>a,b</sup> |  |
| --- | --- | --- | --- | --- |
|  | Difference in means (95% CI) | P Value | Difference in means (95% CI) | P Value |
| PHQ-9 scores<sup>a</sup> | -0.34 (-1.50, 0.83) | 0.567 | -0.22 (-1.37, 0.93) | 0.703 |
| GAD-7 scores<sup>a</sup> | -0.25 (-1.33, 0.84) | 0.657 | -0.48 (-1.75, 0.77) | 0.450 |
| EQ-5D-5L scores<sup>a</sup> | 0.020 (-0.003, 0.043) | 0.084 | 0.026 (-0.005, 0.077) | 0.085 |
| ICECAP-O scores<sup>a</sup> | 0.003 (-0.022, 0.029) | 0.804 | 0.002 (-0.024, 0.029) | 0.859 |
| 3-item UCLA scores<sup>a</sup> | 0.22 (-0.07, 0.50) | 0.142 | 0.22 (-0.07, 0.51) | 0.138 |

<sup>a</sup> Differences in means were estimated using linear regression models, adjusted for relevant baseline assessment of corresponding outcome, gender, age-group, baseline severity of PHQ-9 score (categorical)
<sup>b</sup> All estimates had missing data imputed separately, by trial arm, using MICE models that included predictors of missingness, gender, age-group, baseline severity of PHQ-9 score (categorical)
PHQ-9 scores, corresponding baseline value for outcome in question
PHQ-9 scores range from 0 to 27, with higher scores representing more severe symptoms of depression
GAD-7 scores ranges from 0 to 21 with higher scores representing more severe symptoms of anxiety
EQ-5D-5L scores range from -0.264 to 1, with higher scores representing higher quality of life
ICECAP-O scores range from 0 to 1, with higher scores representing greater levels of capability and general wellbeing
3-item UCLA scores range from 3 to 9, with higher scores representing more severe levels of loneliness

### Sensitivity analyses checking for maintenance of randomisation and subgroup analyses testing for moderation (complete cases only)

#### Sensitivity analysis to determine if randomisation was maintained at the three– and five-month follow-up visits

The comparison at the three-month visits (eTable 11) suggests a slight imbalance whereby participants in the intervention arm had higher levels of hypertension and a greater proportion of participants with more severe forms of depression, compared with participants in the control arm. Additionally, at the three-month follow-up visit participants in the control arm received a greater proportion of participants reporting pharmacological treatment for depression and a greater proportion of participants on higher incomes, than participants in the intervention arm. At the five-month follow-up visit (eTable 12), participants in the control arm had higher levels of hypertension and a greater proportion of people on higher incomes, compared to participants in the intervention arm. However, none of these observed differences were deemed to be sufficiently large to warrant adjustment in the relevant secondary analyses.

**eTable 9:** Comparison of baseline demographics and baseline measures for secondary outcomes, between trial arms, for participants followed-up at the three-month follow-up visit.

| Number/total sample size (%) |  |  |
| --- | --- | --- |
|  | Viva Vida (n=257) | Enhanced usual care (n=270) |
| **Female, No. (%)** | 193/257 (75.1) | 198/270 (73.3) |
| **Age group, No. (%)** |  |  |
| 60-69 years | 214/257 (83.3) | 225/270 (83.3) |
| 70+ years | 43/257 (16.7) | 45/270 (16.7) |
| **Education, No. (%)** |  |  |
| None | 39/256 (15.2) | 44/267 (16.5) |
| 1-4 years | 70/256 (27.3) | 77/267 (28.8) |
| 5-8 years | 83/256 (32.4) | 72/267 (27.0) |
| >8 years | 64/256 (25.0) | 74/267 (27.7) |
| **Personal income, No. (%)** |  |  |
| Up to 1 MW | 172/257 (66.9) | 171/270 (63.3) |
| >1-2 MW | 50/257 (19.5) | 47/270 (17.4) |
| >2 MW | 35/257 (13.6) | 52/270 (19.3) |
| **Hypertension (self-reported), No (%)** | 178 (69.3) | 202/270 (74.8) |
| **Diabetes (self-reported), No (%)** | 106/257 (41.3) | 108/270 (40.0) |
| **Receiving pharmacological treatment for depression (self-reported), No (%)** | 33/257 (12.8) | 41/270 (15.2) |
| **PHQ-9 severity group, No. (%)** |  |  |
| 10-14 | 100/257 (38.9) | 105/270 (38.9) |
| 15-19 | 91/257 (35.4) | 88/270 (32.6) |
| 20+ | 66/257 (25.7) | 77/270 (28.5) |

Abbreviations: MW: minimum wage (in 2021, the minimum wage in Brazil was BRL1110 (approximately US\$213); PHQ-9: 9-item Patient Health Questionnaire for depression.

**eTable 10:** Comparison of baseline demographics and secondary outcome measures, between trial arms, for participants followed-up at the five-month follow-up visit.

| Number/total sample size (%) |  |  |
| --- | --- | --- |
| Baseline variables | Viva Vida (n=250) | Enhanced usual care (n=260) |
| **Female, No. (%)** | 186/250 (74.4) | 188/260 (72.3) |
| **Age group, No. (%)** |  |  |
| 60-69 years | 210/250 (84.0) | 216/260 (83.1) |
| 70+ years | 40/250 (16.0) | 44/260 (16.9) |
| **Education, No. (%)** |  |  |
| None | 38/249 (15.3) | 41/257 (16.0) |
| 1-4 years | 63/249 (25.3) | 75/257 (29.2) |
| 5-8 years | 81/249 (32.5) | 70/257 (27.2) |
| >8 years | 67/249 (26.9) | 71/257 (27.6) |
| **Personal income, No. (%)** |  |  |
| Up to 1 MW | 163/250 (65.2) | 162/260 (62.3) |
| >1-2 MW | 50/250 (20.0) | 46/260 (17.7) |
| >2 MW | 37/250 (14.8) | 52/260 (20.0) |
| **Hypertension (self-reported), No (%)** | 168/250 (67.2) | 193/260 (74.2) |
| **Diabetes (self-reported), No (%)** | 106/250 (42.4) | 102/260 (39.2) |
| **Receiving pharmacological treatment for depression (self-reported), No (%)** | 33/250 (13.2) | 39/260 (15.0) |
| **PHQ-9 severity group, No. (%)** |  |  |
| 10-14 | 102/250 (40.8) | 104/260 (40.0) |
| 15-19 | 87/250 (34.8) | 87/260 (33.5) |
| 20+ | 61/250 (24.4) | 69/260 (26.5) |

Abbreviations: MW: minimum wage (in 2021, the minimum wage in Brazil was BRL 1110 (approximately US\$213); PHQ-9: 9-item Patient Health Questionnaire for depression.

#### Moderation by pre-specified variables for either primary or secondary outcomes

None of the pre-specified subgroup analyses performed to investigate the potential differential intervention effects on the primary outcome of recovery was found to be relevant based on the significance of the corresponding likelihood ratio tests. eTable 13 shows the results of the likelihood ratio statistic for models with and without the interaction term at three months, and eTable 14 shows the results at five months. Multiplicative interactions were assessed on the log odds scale.

**eTable 11:**
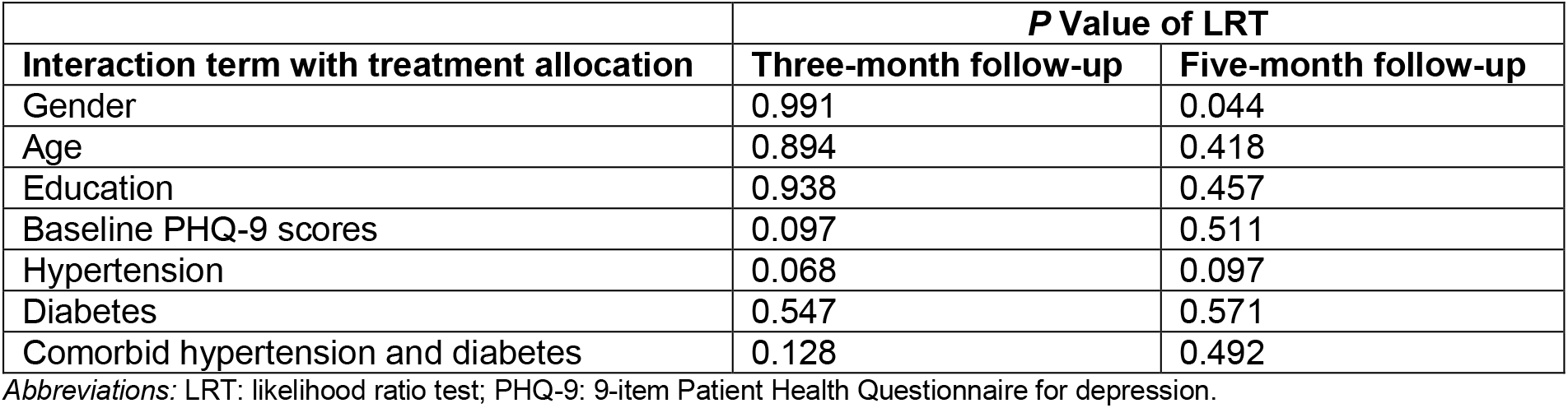
Results of likelihood ratio tests for models with and without the interaction term at three and five months for the primary outcome of recovery from depression.

| Interaction term with treatment allocation | P Value of LRT |  |
| --- | --- | --- |
|  | Three-month follow-up | Five-month follow-up |
| Gender | 0.991 | 0.044 |
| Age | 0.894 | 0.418 |
| Education | 0.938 | 0.457 |
| Baseline PHQ-9 scores | 0.097 | 0.511 |
| Hypertension | 0.068 | 0.097 |
| Diabetes | 0.547 | 0.571 |
| Comorbid hypertension and diabetes | 0.128 | 0.492 |

Abbreviations: LRT: likelihood ratio test; PHQ-9: 9-item Patient Health Questionnaire for depression.

#### Sensitivity analyses checking for potential effect between mean number of elapsed days between baseline and the first follow-up visit

An additional analysis was performed to explore the potential effect of differences in the mean number of days elapsed from baseline to the first follow-up between the intervention arm (median number of days (MDN)=88, intra quartile range (IQR) 87-93) and control arm (MDN=88, IQR 87-90). Estimates from models using complete cases only, that adjusted for mean number of days, were very similar (Odds ratio for difference in recovery from depression at three months between intervention and control arm: 1.54; 95% CI: 1.05, 2.24), to the model that did not adjust for mean number of days (1.58; 1.09, 2.29).

